# Digital Assessment of Objective and Patient-Reported Cognition Across Migraine Phases: Results from the MIND Cohort

**DOI:** 10.64898/2026.04.14.26350892

**Authors:** Babak Khorsand, Devin Teichrow, Crystal J. Jicha, Mia T. Minen, Elizabeth Seng, Richard B. Lipton, Ali Ezzati

## Abstract

**Objective:** Migraine attacks are frequently accompanied by patient-reported “subjective” cognitive symptoms, but objective findings have been inconsistent. We used high-frequency, smartphone-based cognitive testing to assess within-person changes in subjective and objective cognition across migraine phases using daily diaries.

**Methods:** Adults with migraine were recruited through social media. Eligible participants met ICHD-3 migraine criteria and had 3 to 22 monthly headache days. For 30 days, they completed daily smartphone-based reports on headache features, cognitive symptoms, and three smartphone-based objective cognitive tasks. Objective tests included Symbol Search (processing speed/visual search), Color Dots (visual working memory/attention), and Grid Memory (visuospatial working memory). Primary analyses contrasted assessments on current headache days (ictal) versus days with no headache (nonictal). When possible, non-ictal days were subclassified using information from adjacent days as pre-ictal, post-ictal, and interictal days. Outcomes included subjective cognition, reaction time (mean across correctly scored trials), accuracy, and a speed-accuracy composite (Reaction Time/Accuracy). Mixed-effects models adjusted for age, sex, and practice effects.

**Results:** The 139 eligible participants (84.9% female; mean age 38.2 years) contributed 3,014 person-days for ictal versus nonictal comparisons (2,097 nonictal; 917 ictal); for 1,828 person-days precise phase classification was possible. Subjective cognitive symptoms were worse on ictal days, with higher odds of more severe brain fog (OR=3.39, 95% CI 2.70-4.27) and task forgetting (OR=2.82, 95% CI 2.29-3.49). In adjusted models, reaction times were slower on ictal days for Symbol Search (reaction time ratio =1.043, 95% CI 1.028-1.059) and Color Dots (ratio=1.015, 95% CI 1.003-1.026) but not Grid Memory (reaction time ratio =1.006, 95% CI 0.985-1.028). Grid Memory accuracy was lower on ictal days (OR=0.867, 95% CI 0.823-0.914). In analyses based on phase, most nonictal phases showed faster reaction time and lower subjective symptom burden relative to ictal days, with limited differentiation among preictal, postictal, and interictal periods.

**Conclusions:** In persons with migraine, daily smartphone assessments revealed subjective cognitive impairment on ictal vs nonictal days in brain fog and forgetfulness. Objective testing revealed slowing in processing speed and attention and modest differences in the accuracy of working-memory. High-frequency digital cognition appears feasible and may provide scalable functional endpoints for real-world monitoring and treatment evaluation.

## INTRODUCTION

Migraine is a leading cause of a leading cause of years lived with disability worldwide, its burden extends well beyond headache to include fatigue, sensory sensitivity, mood changes, and cognitive dysfunction ^1^. Cognitive symptoms, often described as “brain fog,” slowed thinking, and difficulty concentrating, are common are common and clinically meaningful features of migraine and have been well documented during attacks. ^2, 3^ These symptoms may disrupt daily functioning, self-management, and workplace productivity ^4-6^. Large observational studies have shown that up to half of individuals with migraine report subjective cognitive complaints, which correlate with greater attack frequency, mood disturbance, and sleep disruption^7-9^. Registry and specialty-clinic data likewise suggest that cognitive dysfunction is a meaningful component of migraine burden^10^. Despite their clinical salience, cognitive symptoms are rarely quantified as measurable outcomes in routine care or therapeutic trials.

Studies using objective cognitive testing generally show small or no cognitive differences between persons with migraine and controls^11^. Results vary across studies due in part to differences in participant recruitment, study design, choice of control groups, and choice of cognitive tests. An important source of this heterogeneity is the timing of cognitive assessment relative to the migraine cycle, because deficits are more likely to be detected during ictal or peri-ictal periods than during interictal testing, where findings are often smaller and less consistent. ^2, 12^ This is particularly relevant because subjective cognitive deficits reported by people with migraine also vary across phases ^2, 12^. The relative paucity of objective findings is sometimes used to undermine the credibility of subjective reports of cognitive impairment. Additionally, subjective cognitive impairment may reflect factors beyond objective impairment itself, such as fatigue or avoidance of cognitive exertion due to the belief that mental effort is a migraine trigger ^13^. Clarifying the timing, nature, and determinants of migraine-related cognitive dysfunction is therefore an important step toward making cognition a credible treatment target.

One reason objective findings have been inconsistent is that subjective reports and objective cognitive tests often capture different aspects of migraine-related cognitive dysfunction. Subjective reports reflect a within-person comparison: the patient with migraine reports that, relative to their headache-free state, they experience brain fog or difficulty concentrating or remembering during migraine. In contrast, objective assessments have more often relied on between-group comparisons. This mismatch in design may obscure clinically meaningful phase-dependent changes, particularly because in the interictal state there is little consistent evidence that people with migraine have impaired cognition relative to migraine-free controls with similar demographic characteristics ^11^.

Objective measurement during the ictal phase is rare because bringing patients into clinic during an attack is difficult and repeated in-person testing is limited by burden, cost, and retest effects. Advances in digital testing make frequent home-based assessment feasible and provide a practical way to address these challenges ^14, 15^. In preliminary ambulatory work, repeated smartphone-based cognitive testing detected differences between ictal and nonictal migraine phases^16^. However, it remains unknown whether these within-person changes can be characterized in a larger cohort, across multiple cognitive measures, and throughout the migraine cycle.

To address this gap, we conducted this prospective, longitudinal observational study, MIND (Migraine Impact on Neurocognitive Dynamics). Using daily smartphone-based assessments over 30 days, we collected information on headache characteristics, subjective cognition, and objective cognition in real time. We sought to quantify within-person differences in subjective reports and objective performance, measured by reaction time and accuracy, first by contrasting headache days with all other days and then by examining variation across preictal, ictal, postictal, and interictal phases. Subjective cognitive dysfunction was assessed using daily ratings of brain fog and same-day forgetfulness. In parallel, objective cognitive performance was evaluated using brief smartphone-based tasks spanning multiple cognitive domains, with a focus on cognitive efficiency and potential speed-accuracy tradeoffs. We hypothesized that subjective cognitive dysfunction would peak during ictal periods, with intermediate impairment in the preictal and postictal periods, and that objective performance would also be worse on ictal days, with the largest effects on cognitive efficiency.

## METHODS

### Study Design and Setting

This prospective, decentralized, longitudinal observational study examined within-person changes in subjective and objective cognition across migraine phases in daily life. Participants completed daily smartphone-based symptom diaries and cognitive testing for 30 consecutive days.

### Participants, recruitment, and eligibility

Recruitment occurred between February 2024 and August 2025 via social media platforms such as Reddit and through a small number of local clinic referrals (N=10). Migraine diagnosis was confirmed during screening using International Classification of Headache Disorders, 3rd edition (ICHD-3) criteria and an AMPP-based symptom assessment module to ensure consistency with established population-based migraine methodology ^17^.

Eligibility criteria included age 18 to 65 years, ownership of a compatible smartphone, English proficiency sufficient to complete app-based surveys and tasks, ICHD-3 migraine diagnosis confirmed at screening based on AMPP criteria, and self-reported headache frequency of 3 to 22 headache days per month at screening to increase the likelihood of capturing both headache and headache-free days during the 30-day monitoring period. Exclusion criteria included neurologic disorders other than migraine, history of traumatic brain injury, severe psychiatric illness that would interfere with participation, and current alcohol or drug abuse, as assessed during screening.

### Study procedures

At enrollment, participants completed a comprehensive baseline smartphone-based survey assessing demographics and clinical characteristics. Baseline domains included sociodemographic characteristics (age, sex, race/ethnicity, education, and employment); migraine characteristics (ICHD-3 subtype, attack frequency, typical duration, and pain features); disability and burden measures, including the Migraine Disability Assessment Scale (MIDAS) ^18^, Migraine Severity Scale Score (MSSS) ^19^, and Migraine Treatment Optimization Questionnaire-6 (mTOQ-6) ^17^, Migraine Interictal Burden Scale (MIBS) ^20^, and affective and stress measures, including the Patient Health Questionnaire-4 (PHQ-4) ^21^ and Perceived Stress Scale-4 (PSS-4) ^22^. Baseline measures were used to characterize the cohort (Table 1) and, where applicable, were included as covariates or used for stratified and secondary analyses.

**Table 1.** Baseline cohort characteristics (participant-level)

| Characteristic | Overall (N=139) | Female (n=118) | Male (n=21) |
| --- | --- | --- | --- |
| Age | 38.18 (11.38) | 38.08 (11.02) | 38.71 (13.50) |
| Education: > high school, n (%) | 135 (97.1) | 114 (96.6) | 21 (100.0) |
| Employed (full/part-time), n (%) | 99 (71.2) | 81 (68.6) | 18 (85.7) |
| Hispanic ethnicity: yes, n (%) | 8 (5.8) | 7 (5.9) | 1 (4.8) |
| Race: White only, n (%) | 117 (84.2) | 102 (86.4) | 15 (71.4) |
| Self-reported monthly headache days | 13.76 (6.96) | 13.92 (6.76) | 12.86 (8.12) |
| MSSS | 25.29 (2.51) | 25.66 (2.08) | 23.24 (3.58) |
| MIDAS | 74.68 (64.72) | 77.53 (66.49) | 59.05 (52.57) |
| MIBS | 15.58 (3.62) | 15.89 (3.49) | 13.81 (3.93) |
| Completed study days (out of 30) | 22.26 (7.02) | 22.24 (6.92) | 22.36 (7.69) |
| Captured headache days during study | 6.77 (3.95) | 6.88 (3.97) | 6.18 (3.90) |

**Footnote.** Values are mean (SD) or n (%). Percentages use the full column denominators. Missing data (overall): education n=1, employment n=2, Hispanic ethnicity n=1, race n=2. MSSS = Migraine Symptom Severity Score; MIDAS = Migraine Disability Assessment; MIBS = Migraine Interictal Burden Scale; MHD = monthly headache days.

For 30 consecutive days, participants completed one daily migraine diary and one daily cognitive task battery within a study app developed by our team on the MetricWire platform. Participants received an initial prompt at 12:00 pm local device time and, if incomplete, hourly reminders until 11:59 pm. Assessments not completed by midnight pm were treated as missed for that day. All data were transmitted automatically and stored on secure study servers.

### Analysis sample and data quality exclusions

A total of 186 participants enrolled. For analytic inclusion, participants were required to contribute at least two ictal days and at least two nonictal days during the 30-day period (n = 150). Participants were additionally excluded for persistently low-quality cognitive task data, defined as mean accuracy more than 3 standard deviations below the sample mean (quality-control exclusion; see Supplementary Methods for the operational definition). The final analytic sample included 139 participants (Figure 1).

**Figure 1.**
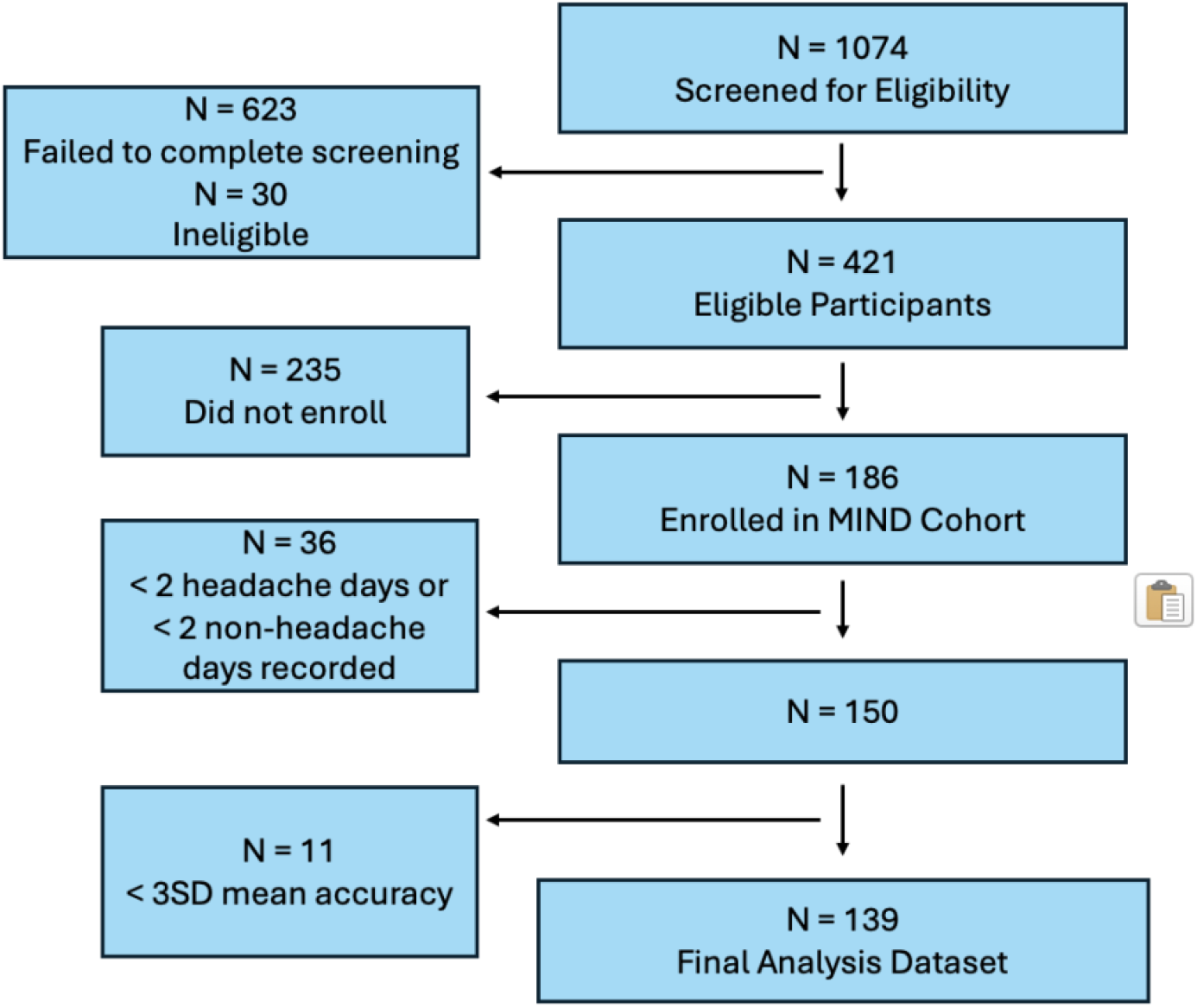
Flowchart of study participants.

### Daily diary measures and migraine phase definitions

Migraine phase was derived from two daily diary items: “Do you have a headache right now?” and “Since the last assessment, did you have a headache?” For the primary contrast (current headache vs no current headache), a person-day was classified as ictal when the participant responded “yes” to the current headache item; days with “no” were classified as nonictal. This definition maximized usable person-days because it relied only on same-day information at the time of assessment.

For phase-resolved analyses, person-days were further classified using adjacent-day diary information into four phases: ictal (current headache present), preictal (a headache-free day immediately preceding an ictal day), postictal (a headache-free day immediately following an ictal day), and interictal (a headache-free day not adjacent to an ictal day). The “since the last assessment” item was used to increase specificity by confirming intervening headache activity and reducing ambiguity around onset and offset between assessments. When adjacent-day diary entries were missing, the person-day was retained for the primary current headache vs no current headache analysis but was not assigned a phase label for preictal, postictal, or interictal analyses.

### Subjective Cognitive outcomes

Subjective cognitive symptoms were assessed daily using two patient-reported items collected alongside the cognitive tasks. Brain fog was measured with the question, “Did you feel that you have a ‘brain fog’ and are not as sharp as usual today?” and rated on a 5-point Likert scale, with higher values indicating greater severity. Forgetfulness was assessed with the question, “Did you forget any tasks today?” with a binary yes/no response. For descriptive tables, brain fog was dichotomized as moderate-to-severe (ratings 3-5), whereas primary models treated brain fog as an ordinal outcome.

### Objective Cognitive Outcomes

Three cognitive tests from the Mobile Monitoring of Cognitive Change platform were administered once daily, immediately after administration of daily surveys: Symbol Search, Color Dots, and Grid Memory (Figure 2) ^14, 23^. Symbol Search is a speeded forced-choice visual search task assessing processing speed and attention; Color Dots is a cued-recall visual working memory binding task requiring recall of color and location information; and Grid Memory is a delayed free-recall visuospatial working memory task requiring reproduction of dot locations on a grid. Each session included 15 Symbol Search trials and 6 trials each for Color Dots and Grid Memory.

**Figure 2.**
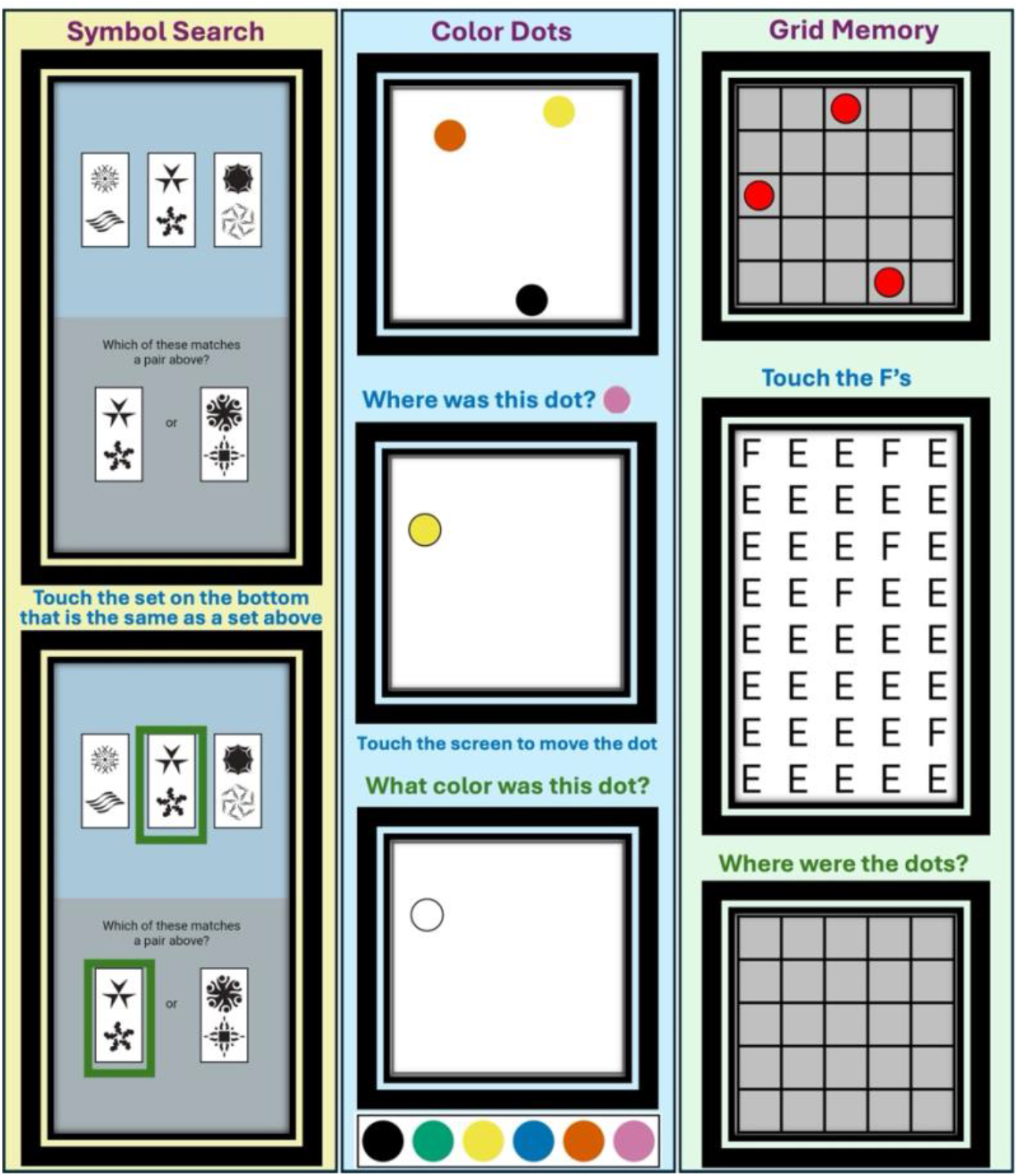
Smartphone-based cognitive tasks used for daily assessment. Symbol Search (processing speed, speeded visual search/attention) is a forced-choice matching task in which participants select the option that matches one of the target tiles. Color Dots (visual working memory binding) requires encoding colored dot locations and then responding to prompts about dot location and color following a brief delay. Grid Memory (visuospatial working memory) requires encoding dot locations on a grid, completing a brief distraction step (illustrated by the “Touch the F’s” screen), and then reproducing the dot locations. Screenshots are representative; specific stimuli, locations, and colors vary across trials. Each daily session included 15 Symbol Search trials and 6 trials each for Color Dots and Grid Memory.

Trial-level reaction time and accuracy were recorded and summarized at the daily assessment level. The primary objective outcome for all tasks was reaction time, defined as the mean reaction time across correctly scored trials (ms), with higher values indicating slower performance. Secondary objective outcomes included (1) accuracy, summarized as the proportion of trials answered correctly each day (0 to 1), and (2) a speed-accuracy composite computed as reaction time/accuracy, with higher values indicating worse performance. As an alternate reaction time definition, we also computed mean reaction time across all trials, including incorrect trials.

### Statistical Analysis

All analyses were conducted at the person-day level, with repeated daily observations nested within participants. We report results using two complementary contrasts: (1) current headache versus no current headache at the time of assessment and (2) four-phase analyses (preictal, ictal, postictal, interictal) derived using adjacent-day diary information. Because phase classification required surrounding diary data, four-phase analyses were restricted to the subset of person-days with sufficient adjacent-day information; sample sizes for each contrast are reported in the Results.

For each cognitive task, person-day outcomes were modeled using mixed-effects models to account for within-person correlation across repeated measures. Reaction time and the composite performance metric (reaction time/accuracy) were positively skewed and analyzed using linear mixed-effects models with log-transformed outcomes. Models included migraine phase (binary or four-level), age, sex, and log(day in study) to account for retest effects over the 30-day protocol, with participant-specific random intercepts and random slopes for log(day) where supported. Exponentiated coefficients from log-transformed models are reported as ratios; percent differences were calculated as (ratio - 1) × 100.

Accuracy (proportion correct) was modeled using generalized mixed-effects models with a binomial distribution and logit link, including the same fixed effects - migraine phase, age, sex, and log(day in study) - and participant-level random effects. Model diagnostics were evaluated for all mixed models; when convergence or identifiability issues were present, continuous predictors were centered and scaled and, if needed, the random-effects structure was simplified in a prespecified manner (e.g., random intercept only) and applied consistently within an outcome family.

Brain fog severity (5-point item) was analyzed using an ordinal mixed-effects model with migraine phase, age, sex, and log(day in study) as fixed effects and participant-level random intercepts. Forgetfulness (yes/no) was analyzed using logistic mixed-effects models with the same fixed effects and participant-level random intercepts.

Missing diary and cognitive-task data were not imputed. Mixed-effects models used all available observations under a missing-at-random assumption. Statistical tests were two-sided with an alpha level of 0.05, and effect estimates are reported with 95% confidence intervals. All model diagnostics were reviewed to assess assumptions and overall fit, including residual distributions for linear mixed models and convergence diagnostics for generalized and ordinal mixed models. All analyses were conducted in R (version 4.5.0).

### Ethical Considerations

The study protocol was approved by the University of California, Irvine Institutional Review Board. All participants provided informed consent electronically before enrollment. Participants were compensated for their time based on study completion, in accordance with the approved protocol.

### Data Availability

De-identified data are available from the corresponding author upon reasonable request and subject to institutional data-sharing agreements.

## RESULTS

### Study sample and observation counts

A total of 186 participants enrolled; 150 met the minimum data requirement of at least 2 ictal and 2 nonictal days, and after quality-control exclusions the final analytic sample was 139 (Figure 1). The cohort had a mean age of 38.18 years (SD 11.38) and was 84.9% female (118/139), with a mean of 13.76 self-reported monthly headache days (SD 6.96) at baseline (Table 1). Participants contributed 3,014 person-days for the primary ictal versus nonictal comparison (2,097 nonictal; 917 ictal) and 1,828 phase-classified person-days for preictal, ictal, postictal, and interictal analyses (preictal n=196; ictal n=917; postictal n=267; interictal n=448). Across phase-classified analyses, evaluable person-days were 1,828 for Symbol Search and Color Dots and 1,719 for Grid Memory.

### Subjective cognition

Participants experienced more brain fog on ictal days compared with non-ictal days (60.0% vs. 26.8%). Additionally, task forgetting was higher on ictal days than on non-ictal days (31.8% vs. 17.2%) (Table 2).

**Table 2.** Outcomes by headache presence at the time of assessment (person-days)

| Daily outcome | No current headache<br>(N=2097 person-days) | Current headache<br>(N=917 person-days) |
| --- | --- | --- |
| Brain fog (moderate-to-severe), n (%) | 562 (26.8) | 550 (60.0) |
| Forgetfulness, n (%) | 361 (17.2) | 292 (31.8) |
| Symbol Search reaction time (ms) | 1851.09 (546.81) | 1984.93 (666.71) |
| Color Dots reaction time (ms) | 1817.62 (477.08) | 1920.21 (595.39) |
| Grid Memory reaction time (ms) | 4222.12 (1260.22) | 4248.93 (1310.12) |
| Symbol Search accuracy | 0.97 (0.06) | 0.96 (0.06) |
| Color Dots accuracy | 0.94 (0.11) | 0.94 (0.12) |
| Grid Memory accuracy | 0.85 (0.15) | 0.82 (0.17) |
| Symbol Search performance | 1909.19 (557.02) | 2046.56 (666.84) |
| Color Dots performance | 2084.68 (1193.01) | 2220.38 (1118.92) |
| Grid Memory performance | 5649.09 (2286.45) | 5988.93 (2723.57) |

**Footnote.** Values are mean (SD) unless otherwise stated. Reaction time is in milliseconds and reflects mean reaction time across correctly scored trials (primary definition). Accuracy is the fraction of trials answered correctly (range 0 to 1). Performance is calculated as ReactionTime/Accuracy (higher values indicate worse performance). Brain fog was dichotomized for descriptive tables as moderate-to-severe (ratings of 3–5 on the 5-point scale), while primary models treated brain fog as an ordinal outcome.

Across migraine phases, subjective symptoms were most frequent on ictal days and lower on preictal, postictal, and interictal days (Table 3).

**Table 3.** Daily outcomes across 4 migraine phases (person-days)

| Daily outcome | Preictal<br>(N=196) | Ictal<br>(N=917) | Postictal<br>(N=267) | Interictal<br>(N=448) |
| --- | --- | --- | --- | --- |
| Brain fog (moderate-to-severe), n (%) | 58 (29.6) | 550 (60.0) | 53 (19.9) | 77 (17.2) |
| Forgetfulness, n (%) | 39 (19.9) | 292 (31.8) | 28 (10.5) | 53 (11.8) |
| Symbol Search reaction time (ms) | 1865.61<br>(541.72) | 1984.93<br>(666.71) | 1840.85<br>(575.21) | 1831.62<br>(459.59) |
| Color Dots reaction time (ms) | 1835.44<br>(490.80) | 1920.21<br>(595.39) | 1824.18<br>(468.25) | 1780.32<br>(412.39) |
| Grid Memory reaction time (ms) | 4168.96<br>(1232.09) | 4248.93<br>(1310.12) | 4183.28<br>(1209.62) | 4106.49<br>(1185.07) |
| Symbol Search accuracy | 0.97 (0.06) | 0.96 (0.06) | 0.96 (0.08) | 0.97 (0.05) |
| Color Dots accuracy | 0.93 (0.12) | 0.94 (0.12) | 0.93 (0.11) | 0.96 (0.10) |
| Grid Memory accuracy | 0.86 (0.15) | 0.82 (0.17) | 0.85 (0.14) | 0.85 (0.15) |
| Symbol Search performance | 1925.22<br>(550.03) | 2046.56<br>(666.84) | 1908.79<br>(567.26) | 1880.72<br>(467.83) |
| Color Dots performance | 2134.22<br>(1041.83) | 2220.38<br>(1118.92) | 2107.21<br>(967.44) | 1954.03<br>(690.51) |
| Grid Memory performance | 5493.84<br>(2235.97) | 5988.93<br>(2723.57) | 5642.83<br>(2152.05) | 5527.48<br>(2281.10) |

**Footnote:** Values are mean (SD) unless otherwise stated. Reaction time is in milliseconds and reflects mean reaction time across correctly scored trials (primary definition). Accuracy is the fraction of trials answered correctly (range 0 to 1). Performance is calculated as ReactionTime/Accuracy (higher values indicate worse performance). Brain fog was dichotomized for descriptive tables as moderate-to-severe (ratings of 3–5 on the 5-point scale), while primary models treated brain fog as an ordinal outcome.

In adjusted mixed-effects models, ictal days were associated with higher odds of more severe brain fog (OR 3.39, 95% CI 2.70-4.27) and task forgetting (OR 2.83, 95% CI 2.29-3.49). In phase-resolved models with ictal days as the reference, odds of both symptoms were lower on preictal, postictal, and interictal days: brain fog OR 0.28 (95% CI 0.17-0.46), 0.19 (0.11-0.31), and 0.16 (0.10-0.25), respectively; forgetfulness OR 0.43 (0.28-0.66), 0.17 (0.11-0.28), and 0.21 (0.14-0.31), respectively (all p<0.001) (Table 4). These results indicate a clear ictal increase in subjective cognitive burden.

**Table 4.** Mixed-effects models for subjective cognitive outcomes across migraine phases (reference = ictal)

| Phase comparison (vs ictal) | Brain fog OR (95% CI) | p | Forgetfulness OR (95% CI) | p |
| --- | --- | --- | --- | --- |
| Preictal vs ictal | 0.28 (0.17–0.46) | <0.001 | 0.43 (0.28–0.66) | <0.001 |
| Postictal vs ictal | 0.19 (0.11–0.31) | <0.001 | 0.17 (0.11–0.28) | <0.001 |
| Interictal vs ictal | 0.16 (0.10–0.25) | <0.001 | 0.21 (0.14–0.31) | <0.001 |

**Footnote:** Values are adjusted odds ratios with 95% confidence intervals from mixed-effects models. Brain fog was modeled using an ordinal mixed-effects model; forgetfulness was modeled using a logistic mixed-effects model. Models adjusted for age, sex, and log(day in study), with participant random effects as specified in Methods. Phase-resolved models were restricted to person-days with sufficient adjacent-day diary data for classification.

### Objective cognition

Daily descriptive outcomes by headache status are summarized in Table 2. Ictal days were characterized by slower reaction times on Symbol Search (1985 ± 667 ms vs 1851 ± 547 ms) and Color Dots (1920 ± 595 ms vs 1818 ± 477 ms) compared with nonictal days, while Grid Memory reaction time was similar (4249 ± 1310 ms vs 4222 ± 1260 ms). Grid Memory accuracy was lower on ictal days (0.82 ± 0.17 vs 0.85 ± 0.15). Daily outcomes across the 4 migraine phases are summarized in Table 3.

In adjusted mixed-effects models, ictal days were associated with slower reaction time for Symbol Search (reaction time ratio 1.043, 95% CI 1.028-1.059, p<0.001) and Color Dots (1.015, 95% CI 1.003-1.026, p=0.013), but not for Grid Memory (1.006, 95% CI 0.985-1.028, p=0.563). Grid Memory accuracy was reduced on ictal days (OR 0.867, 95% CI 0.823-0.914, p<0.001). For the speed-accuracy composite, ictal days were associated with worse performance on Symbol Search (performance ratio 1.043, 95% CI 1.028-1.058, p<0.001) and Grid Memory (1.037, 95% CI 1.014-1.061, p=0.001), with weaker evidence for Color Dots (1.017, 95% CI 0.998-1.037, p=0.085) (Table 5). Results were similar using the alternate reaction-time definition that included all trials (Table S1).

**Table 5.**
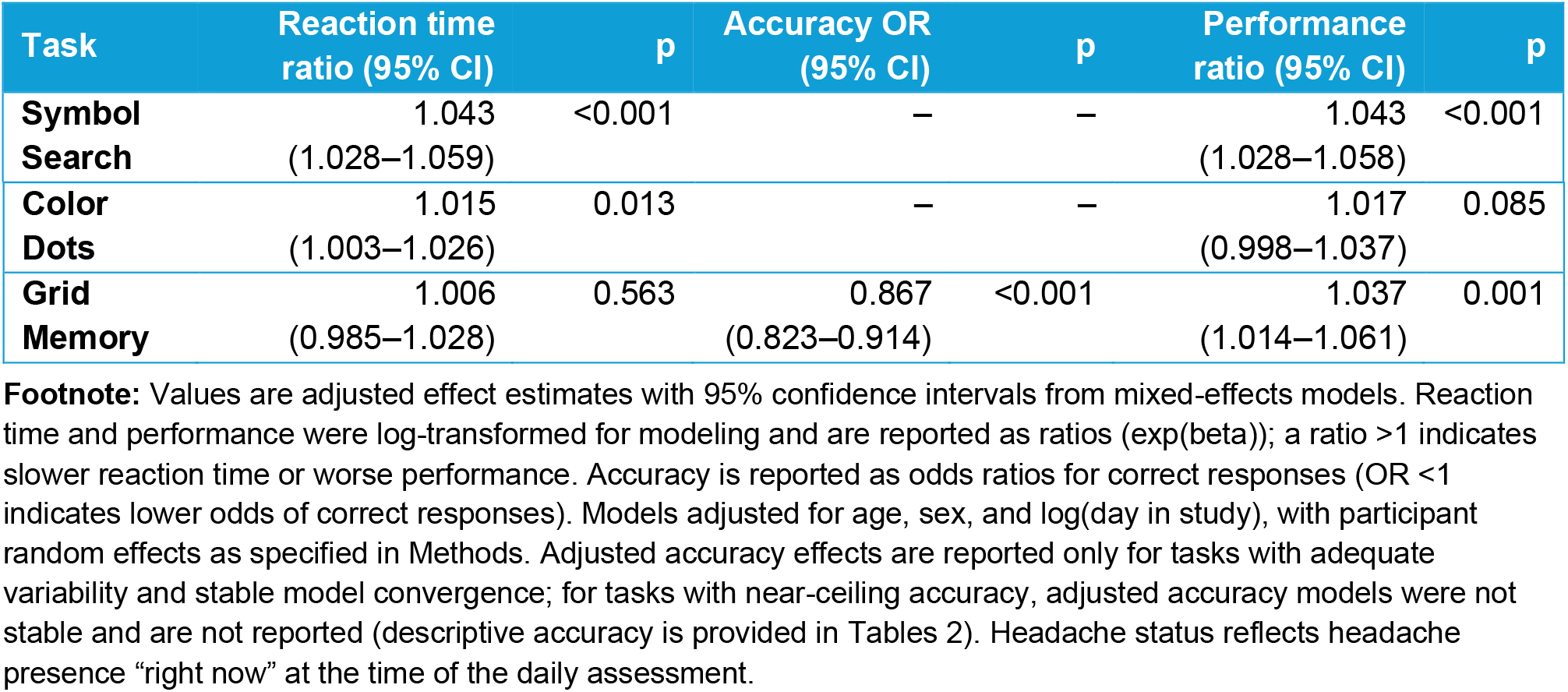
Mixed-effects models for objective cognitive outcomes by headache presence at the time of assessment.

In phase-resolved models with ictal days as the reference, reaction times were faster during nonictal phases, with the most consistent differences observed for Symbol Search: preictal 0.954 (95% CI 0.925-0.984, p=0.003), postictal 0.933 (0.909-0.959, p<0.001), and interictal 0.955 (0.931-0.979, p<0.001). For Color Dots, reaction time was faster postictally (0.968, 95% CI 0.949-0.986, p<0.001), whereas preictal (0.984, 0.963-1.006, p=0.147) and interictal (0.984, 0.966-1.002, p=0.074) differences were smaller. For Grid Memory, reaction time was faster interictally (0.958, 95% CI 0.926-0.990, p=0.012), with no significant differences preictally (0.989, 0.949-1.031, p=0.608) or postictally (0.979, 0.944-1.016, p=0.260). Grid Memory accuracy was higher on preictal (OR 1.370, 95% CI 1.231-1.525, p<0.001), postictal (1.231, 1.123-1.349, p<0.001), and interictal days (1.206, 1.106-1.316, p<0.001) than on ictal days (Table 6). Figure 3 shows the model-estimated phase patterns with 95% CIs.

**Table 6.** Mixed-effects models for objective cognitive outcomes across migraine phases (reference = ictal)

| Task and phase comparison (vs ictal) | Reaction time ratio (95% CI) | p | Accuracy OR (95% CI) | p | Performance ratio (95% CI) | p |
| --- | --- | --- | --- | --- | --- | --- |
| Symbol Search - preictal vs ictal | 0.954<br>(0.925–0.984) | 0.003 | — | — | 0.954<br>(0.926–0.982) | 0.002 |
| Symbol Search - postictal vs ictal | 0.933<br>(0.909–0.959) | <0.001 | — | — | 0.940<br>(0.917–0.965) | <0.001 |
| Symbol Search - interictal vs ictal | 0.955<br>(0.931–0.979) | <0.001 | — | — | 0.949<br>(0.927–0.972) | <0.001 |
| Color Dots - preictal vs ictal | 0.984<br>(0.963–1.006) | 0.147 | — | — | 0.986<br>(0.951–1.022) | 0.446 |
| Color Dots - postictal vs ictal | 0.968<br>(0.949–0.986) | <0.001 | — | — | 0.970<br>(0.940–1.001) | 0.059 |
| Color Dots - interictal vs ictal | 0.984<br>(0.966–1.002) | 0.074 | — | — | 0.967<br>(0.939–0.996) | 0.027 |
| Grid Memory - preictal vs ictal | 0.989<br>(0.949–1.031) | 0.608 | 1.370<br>(1.231–1.525) | <0.001 | 0.931<br>(0.892–0.972) | 0.001 |
| Grid Memory - postictal vs ictal | 0.979<br>(0.944–1.016) | 0.260 | 1.231<br>(1.123–1.349) | <0.001 | 0.963<br>(0.928–1.000) | 0.053 |
| Grid Memory - interictal vs ictal | 0.958<br>(0.926–0.990) | 0.012 | 1.206<br>(1.106–1.316) | <0.001 | 0.933<br>(0.900–0.966) | <0.001 |

**Figure 3.**
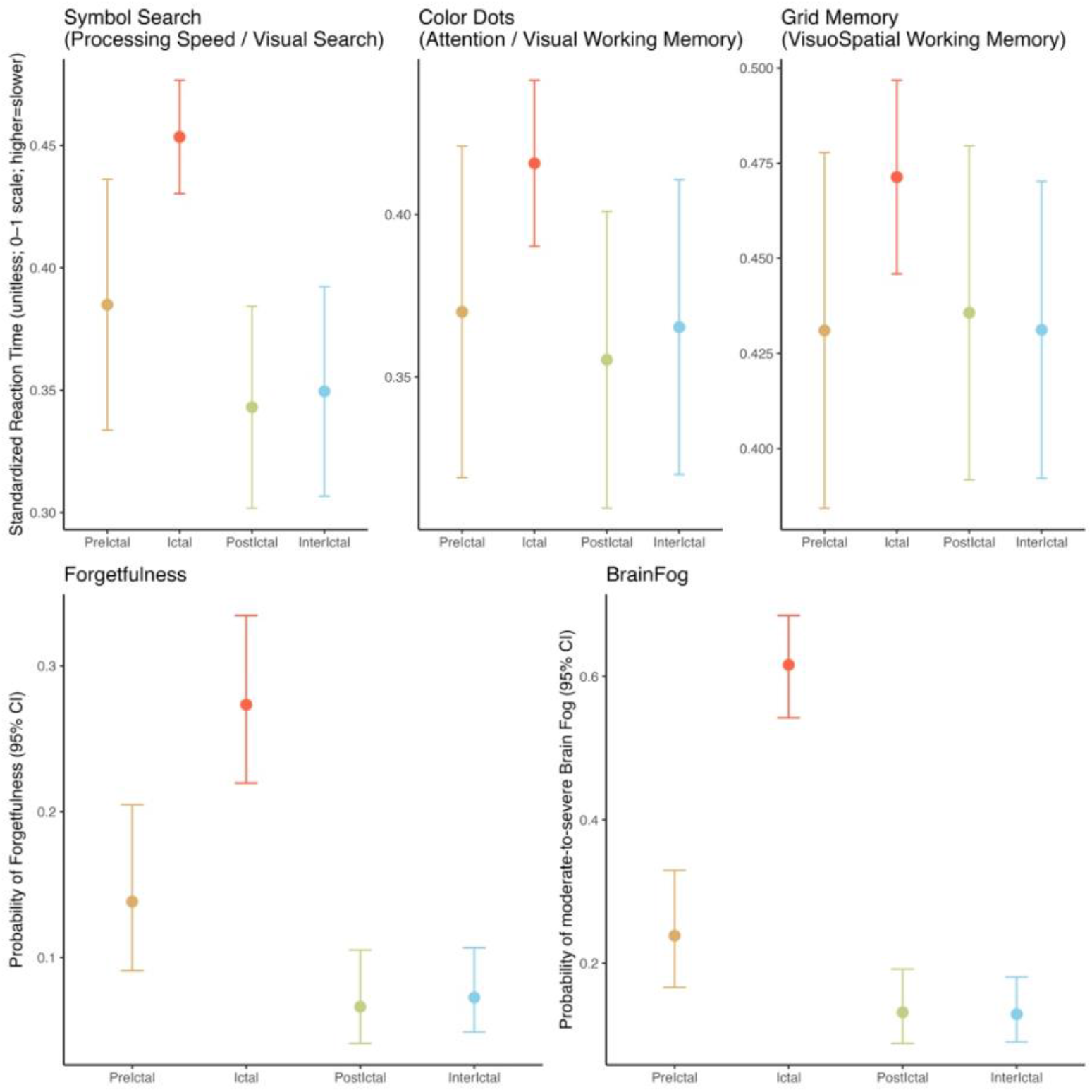
Phase-Dependent Changes in Objective and Patient-Reported Cognition Across the Migraine Cycle. Panels A–C show adjusted mean standardized reaction time (0–1 scale; higher = slower) for Symbol Search, Color Dots, and Grid Memory across migraine phases. Panels D–E show adjusted probabilities of task forgetting and moderate-to-severe brain fog. Points are model-estimated phase-specific effects; error bars indicate 95% CIs. Phases: preictal (day before), ictal (headache day), postictal (day after), interictal.

In supplementary analyses using the alternate all-trials reaction-time definition with interictal as the reference (Table S2), preictal and postictal estimates were generally close to interictal for Symbol Search and Color Dots. For Grid Memory, postictal days showed nominal slowing relative to interictal days (1.038, 95% CI 1.003-1.074, p=0.032). Overall, these supplementary results support clear separation between ictal and nonictal performance, with limited differentiation among nonictal phases.

## DISCUSSION

Daily smartphone-based cognitive testing detected robust, phase-dependent changes in cognition among adults with migraine, with reaction time consistently slowest on ictal days for Symbol Search (+4.3%) and Color Dots (+1.5%). Grid Memory reaction time differences were smaller - the primary ictal vs nonictal contrast was not significant, although interictal days were modestly faster than ictal days in phase-specific analyses (reaction time ratio 0.958). Working-memory accuracy also declined ictally (Grid Memory accuracy OR=0.87), and these objective patterns paralleled substantially higher odds of patient-reported brain fog (OR=3.39) and same-day task forgetting (OR=2.82) during attacks. Together, these findings support high-frequency digital cognitive assessment as a feasible and sensitive approach for quantifying state-dependent cognitive burden in real-world settings. They also support the clinical observation that cognitive symptoms are a frequent and disabling part of migraine, often reported across the premonitory, headache, and postdrome phases, and sometimes persisting between attacks ^2, 24^.

Mobile cognitive assessments may complement patient-reported outcomes by capturing dimensions of migraine-related dysfunction that matter for work and daily life. Daily digital tasks offer a scalable, patient-centered, and objective outcome to evaluate treatment response, particularly for interventions seeking to reduce the cognitive burden that persists beyond pain metrics. The alignment of objective slowing with subjective fog and forgetting supports integrating performance-based and patient-reported measures in both clinical monitoring and trials.

Phase-based analyses reinforced that the primary separation was between ictal vs nonictal assessments. Reaction times were generally faster on preictal, postictal, and interictal days compared with ictal days, but contrasts among the nonictal phases were mostly null, suggesting limited differentiation between preictal, postictal, and interictal periods at a once-daily sampling frequency. This pattern suggests that our design was more effective in distinguishing attack-related cognitive change than in resolving finer-grained differences among nonictal phases. This is consistent with prior work showing that cognitive disruption is most pronounced during attacks, whereas findings outside attacks are more heterogeneous and appear sensitive to study setting, sampling intensity, and measurement approach ^25^.

Our findings align with more than two decades of neuropsychological research demonstrating cognitive changes across migraine phases. Early clinic-based work reported transient declines in attention, short-term memory, and executive function during attacks, with recovery thereafter, while subsequent studies confirmed slowed processing speed and impaired working memory during ictal and early interictal periods ^24, 26^. Controlled within-person clinic studies have also shown ictal decrements in reading/processing speed and in verbal learning and delayed recall, supporting episodic-memory vulnerability during attacks ^26^. At the between-person level, reviews and meta-analyses suggest small-to-moderate average differences for migraine versus non-migraine controls across multiple domains (often largest for processing speed and executive function), but with substantial heterogeneity and mixed evidence for consistent interictal impairment in population-based samples ^24, 25^. Our study extends this literature into daily life using ecological momentary assessment, showing that within-person differences in reaction time - and, to a lesser extent, task accuracy - can be detected outside the clinic. Prior smartphone EMA work used brief cognitive testing multiple times per day over 2–4 weeks and detected ictal decrements in cognitive inhibition and spatial working memory, supporting the view that ambulatory testing can detect state-dependent effects during pain. ^16^ One plausible mechanism is that pain competes for attentional resources, preferentially affecting speeded visual search, attention, and working-memory performance during attacks.

Up to half of individuals with migraine report cognitive complaints linked to attack frequency, mood disturbance, and sleep disruption ^7-9^. Our results corroborate this patient experience by demonstrating much larger phase effects on brain fog and forgetting than on objective accuracy. This dissociation suggests that patient-perceived cognitive burden may reflect a broader syndrome (e.g., slowed thinking, reduced mental efficiency, decision difficulty) that is only partly captured by brief task accuracy, reinforcing the importance of pairing objective and patient-reported measures.

### Limitations and Future directions

Several limitations should be considered. Time-varying factors such as sleep, fatigue, stress, mood, and acute medication timing may influence both migraine status and cognition, though in some cases may lie on the causal pathway. In addition, migraine phase was inferred from once-daily diaries and adjacent-day rules, and our operational phase definitions do not map perfectly onto clinical phase concepts. Because premonitory symptoms can occur up to 48 hours before headache onset and the postdrome may last 1 to 2 days, day-level phase labels may misclassify the true temporal boundaries of attacks and may blur distinctions among preictal, postictal, and interictal periods ^27^. Second, the study relied on remote, unsupervised, smartphone-based assessment. Performance may have been affected by testing context (distraction, lighting, screen use during photophobia) and by the restricted completion window. Time-of-day effects have been shown in smartphone-based cognitive testing, so variation in when participants completed tasks within the 2–10 pm window could add noise or bias phase comparisons if completion timing differed systematically by migraine status ^28^. Third, the cognitive battery emphasized processing speed/attention and visuospatial working memory and did not assess episodic memory, language, and cognitive inhibition/executive control ^26^. Finally, although we followed published task specifications, accuracy was high for some measures. In a relatively young cohort, limited task difficulty can compress the performance range (ceiling effects) and reduce discrimination of subtle within-person change, indicating that future versions may benefit from higher difficulty or adaptive task parameters to better match participant ability ^14^.

Future work should address these limitations by increasing sampling density (for example, multiple short assessments per day) to better capture onset, peak, and recovery trajectories and to improve detection of preictal and postictal effects. Expanding the battery to include episodic memory and inhibition/executive tasks and introducing adaptive difficulty or item-response-based approaches, may improve sensitivity and reduce ceiling effects in younger, high-functioning samples. Finally, future studies should evaluate responsiveness to acute and preventive treatments and establish reliability and clinically interpretable thresholds for change, building on prior psychometric work in ambulatory cognitive assessment.

### Clinical Implications

Cognitive dysfunction is a meaningful component of migraine burden and merits routine assessment alongside pain and associated symptoms. Qualitative studies describe difficulties with attention, processing, language, and executive function during attacks that can impair medication management, decision-making, and communication ^2, 3^. Clinicians can mitigate this by developing interictal action plans, such as written medication instructions, reminders, and involvement of a trusted support person for high-impact attacks, and by documenting migraine phase and timing of the last attack when interpreting cognitive screening or neuropsychological testing. For nonurgent evaluations, clinicians should also consider whether testing performed during ictal or peri-ictal periods may underestimate usual functioning.

Within the framework of clinical-trial methodology, migraine therapeutics are primarily evaluated via patient-reported outcomes (PROs), such as pain freedom for acute trials and reductions in migraine days for preventive studies ^29, 30^. Although essential in the absence of objective biomarkers, these subjective measures are susceptible to recall bias, measurement error, and data missingness—particularly during ictus ^31, 32^. While FDA guidance mandates validated PRO instruments for labeling claims ^33^, objective digital cognitive measures could provide a low-burden, high-frequency complement to traditional diaries. By quantifying functional dimensions often cited as disabling by patients, these metrics offer an assessment of *cognitive efficiency* that may be less vulnerable to some limitations of self-report. Integrating these tools as exploratory endpoints in future trials would allow for a more comprehensive evaluation of therapeutic benefit beyond pain-centric outcomes alone.

## Conclusion

Daily smartphone cognitive testing captured clinically meaningful, migraine-phase changes in processing speed and working memory and tracked parallel spikes in brain fog and forgetfulness on headache days. These low-burden, objective measures can complement symptom diaries, help interpret cognitive complaints in practice, and provide scalable functional endpoints for trials evaluating acute and preventive therapies. These findings support digital cognitive assessment as a practical approach to measuring real-world cognitive burden across the migraine cycle.

## Data Availability Statement

The datasets used and analyzed during the current study are available from the corresponding author upon reasonable request.

---

**Table S1.**
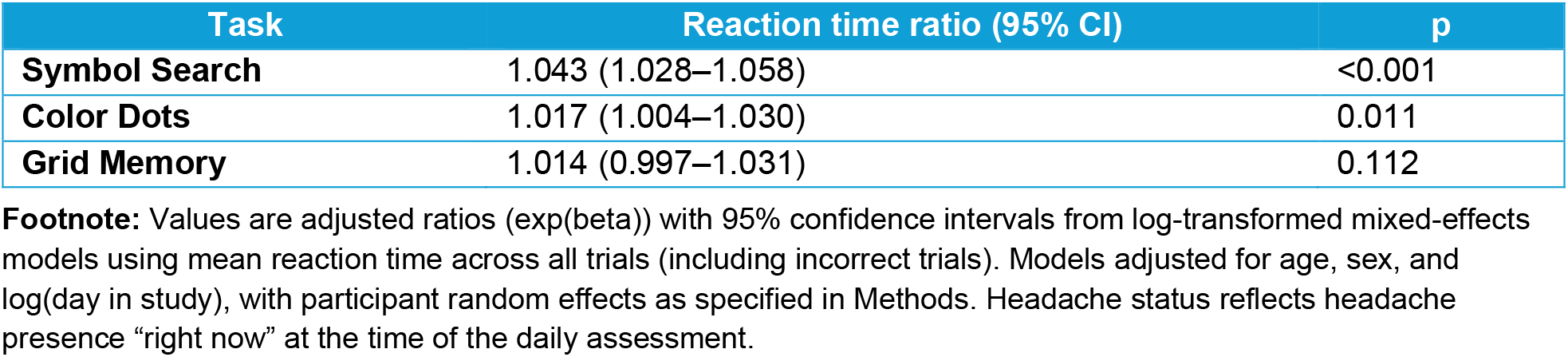
Alternate reaction time definition (all trials) by headache presence at the time of assessment.

| Task | Reaction time ratio (95% CI) | p |
| --- | --- | --- |
| **Symbol Search** | 1.043 (1.028–1.058) | <0.001 |
| **Color Dots** | 1.017 (1.004–1.030) | 0.011 |
| **Grid Memory** | 1.014 (0.997–1.031) | 0.112 |

**Footnote:** Values are adjusted ratios (exp(beta)) with 95% confidence intervals from log-transformed mixed-effects models using mean reaction time across all trials (including incorrect trials). Models adjusted for age, sex, and log(day in study), with participant random effects as specified in Methods. Headache status reflects headache presence “right now” at the time of the daily assessment.

**Table S2.** Alternate reaction time definition (all trials) across migraine phases (reference = interictal)

| Phase comparison (vs interictal) | Symbol Search RT ratio (95% CI), p | Color Dots RT ratio (95% CI), p | Grid Memory RT ratio (95% CI), p |
| --- | --- | --- | --- |
| **Ictal vs interictal** | 1.049 (1.023–1.075), <0.001 | 1.024 (1.004–1.045), 0.019 | 1.044 (1.016–1.072), 0.002 |
| **Preictal vs interictal** | 1.001 (0.969–1.035), 0.934 | 1.006 (0.979–1.034), 0.658 | 1.025 (0.988–1.063), 0.193 |
| **Postictal vs interictal** | 0.982 (0.952–1.012), 0.239 | 1.000 (0.975–1.025), 0.997 | 1.038 (1.003–1.074), 0.032 |

**Footnote:** Values are adjusted ratios (exp(beta)) with 95% confidence intervals from log-transformed mixed-effects models using mean reaction time across all trials (including incorrect trials). Reference phase is interictal. Models adjusted for age, sex, and log(day in study), with participant random effects as specified in Methods. Phase-resolved models were restricted to observations with sufficient adjacent-day diary data for classification.

## References

1. Stovner LJ, Nichols E, Steiner TJ, et al. Global, regional, and national burden of migraine and tension-type headache, 1990–2016: a systematic analysis for the Global Burden of Disease Study 2016. The Lancet Neurology 2018;17:954–976.

2. Gerstein MT, Wirth R, Uzumcu AA, et al. Patient-reported experiences with migraine-related cognitive symptoms: results of the MiCOAS qualitative study. Headache: The Journal of Head and Face Pain 2023;63:441–454.

3. Mangrum R, Bryant AL, Gerstein MT, et al. The impacts of migraine on functioning: results from two qualitative studies of people living with migraine. Headache: The Journal of Head and Face Pain 2024;64:156–171.

4. Begasse de Dhaem O, Robbins MS. Cognitive impairment in primary and secondary headache disorders. Current Pain and Headache Reports 2022;26:391–404.

5. Fernandes C, Dapkute A, Watson E, et al. Migraine and cognitive dysfunction: a narrative review. The Journal of Headache and Pain 2024;25:221.

6. Chu HT, Liang CS, Lee JT, et al. Subjective cognitive complaints and migraine characteristics: a cross-sectional study. Acta Neurologica Scandinavica 2020;141:319–327.

7. Lee SH, Kang Y, Cho S-J. Subjective cognitive decline in patients with migraine and its relationship with depression, anxiety, and sleep quality. The journal of headache and pain 2017;18:77.

8. Lee SH, Cho S-J. Subjective Cognitive Decline Patterns in Patients with Migraine, with or without Depression, versus Non-depressed Older Adults. Headache and Pain Research 2024;25:103–110.

9. Ikechukwu OJ, Khan MN, Khan SM, et al. Association between sleep quality and cognitive complaints among young adults with migraine. Cureus 2025;17.

10. Schwedt TJ, Digre K, Tepper SJ, et al. The American registry for migraine research: research methods and baseline data for an initial patient cohort. Headache: The Journal of Head and Face Pain 2020;60:337–347.

11. Suhr JA, Seng EK. Neuropsychological functioning in migraine: clinical and research implications. Cephalalgia 2012;32:39–54.

12. Braganza DL, Fitzpatrick LE, Nguyen ML, Crowe SF. Interictal cognitive deficits in migraine sufferers: a meta-analysis. Neuropsychology Review 2022;32:736–757.

13. Klepper JE, Sebrow L, Rosen NL, Seng EK. Cogniphobia and neuropsychological functioning in migraine. Neuropsychology 2022;36:433.

14. Sliwinski MJ, Mogle JA, Hyun J, Munoz E, Smyth JM, Lipton RB. Reliability and validity of ambulatory cognitive assessments. Assessment 2018;25:14–30.

15. Oravecz Z, Vandekerckhove J, Hakun JG, et al. Computational phenotyping of cognitive decline with retest learning. The Journals of Gerontology, Series B: Psychological Sciences and Social Sciences 2025;80:gbaf030.

16. Sebrow L, Lipton RB, Metts CL, et al. Cognition during ictal and interictal migraine phases: A pilot study using ecological momentary assessment. Headache: The Journal of Head and Face Pain 2025;65:1684–1692.

17. Lipton RB, Stewart WF, Diamond S, Diamond ML, Reed M. Prevalence and burden of migraine in the United States: data from the American Migraine Study II. Headache: The Journal of Head and Face Pain 2001;41:646–657.

18. Stewart WF, Lipton RB, Dowson AJ, Sawyer J. Development and testing of the Migraine Disability Assessment (MIDAS) Questionnaire to assess headache-related disability. Neurology 2001;56:S20–S28.

19. Lipton RB, Diamond S, Reed M, Diamond ML, Stewart WF. Migraine diagnosis and treatment: results from the American Migraine Study II. Headache: The Journal of Head and Face Pain 2001;41:638–645.

20. Buse DC, Bigal M, Rupnow M, Reed M, Serrano D, Lipton R. Development and validation of the Migraine Interictal Burden Scale (MIBS): a self-administered instrument for measuring the burden of migraine between attacks. Neurology; 2007: LIPPINCOTT WILLIAMS & WILKINS 530 WALNUT ST, PHILADELPHIA, PA 19106-3621 USA: A89–A89.

21. Kroenke K, Spitzer RL, Williams JB, Löwe B. An ultra-brief screening scale for anxiety and depression: the PHQ–4. Psychosomatics 2009;50:613–621.

22. Cohen S, Kamarck T, Mermelstein R. A global measure of perceived stress. Journal of health and social behavior 1983:385–396.

23. Thompson LI, De Vito AN, Kunicki ZJ, et al. Psychometric and adherence considerations for high-frequency, smartphone-based cognitive screening protocols in older adults. Journal of the International Neuropsychological Society 2024;30:785–793.

24. Vuralli D, Ayata C, Bolay H. Cognitive dysfunction and migraine. The journal of headache and pain 2018;19:109.

25. Gu L, Wang Y, Shu H. Association between migraine and cognitive impairment. The journal of headache and pain 2022;23:88.

26. Gil-Gouveia R, Oliveira AG, Martins IP. Cognitive dysfunction during migraine attacks: a study on migraine without aura. Cephalalgia 2015;35:662–674.

27. Karsan N, Goadsby PJ. Biological insights from the premonitory symptoms of migraine. Nature Reviews Neurology 2018;14:699–710.

28. Wilks H, Aschenbrenner AJ, Gordon BA, et al. Sharper in the morning: Cognitive time of day effects revealed with high-frequency smartphone testing. Journal of Clinical and Experimental Neuropsychology 2021;43:825–837.

29. Diener H-C, Tassorelli C, Dodick DW, et al. Guidelines of the International Headache Society for controlled trials of acute treatment of migraine attacks in adults. Cephalalgia 2019;39:687–710.

30. Diener H-C, Tassorelli C, Dodick DW, et al. Guidelines of the International Headache Society for controlled trials of preventive treatment of migraine attacks in episodic migraine in adults. Cephalalgia 2020;40:1026–1044.

31. McKenzie JA, Cutrer FM. How well do headache patients remember? A comparison of self-report measures of headache frequency and severity in patients with migraine. Headache: The Journal of Head and Face Pain 2009;49:669–672.

32. Torelli P, Jensen R. Headache diaries and calendars. Handbook of Clinical Neurology: Elsevier, 2010: 137–146.

33. Varma R, Richman EA, Ferris FL, Bressler NM. Use of patient-reported outcomes in medical product development: a report from the 2009 NEI/FDA Clinical Trial Endpoints Symposium. Investigative ophthalmology & visual science 2010;51:6095–6103.

